# Deep Learning-Based Classification of Bone Lesions on CT Scans of Metastatic Spine Disease Patients: A 3D-Convolutional Neural Network Approach

**DOI:** 10.64898/2026.08.13.26360407

**Authors:** Vy Hong, Bulent Aslan, Nazim Haouchine, Steve Pieper, William Wells, Mario Keko, David Kozono, Patrick F. Doyle, Tracy Balboni, Alexander Spektor, Mai Anh Huynh, David B. Hackney, Ron N. Alkalay

**Author notes:** Address correspondence to: Ron N. Alkalay, PhD, Associate Professor in Orthopedic Surgery, Center for Advanced Orthopedic Studies Beth Israel Deaconess Medical Center, 1 Overland Street, Boston, MA, 02215, USA.

## Abstract

**Purpose:** Clinical assessment of vertebral lesion quality (osteolytic, osteoblastic, mixed) remains subjective, with limited interobserver reliability. This study evaluated a novel application of 3D convolutional neural networks (3D-CNNs) for classifying lesion quality from CT volumes in metastatic cancer patients.

**Materials and Methods:** This retrospective study used CT data from 151 cancer patients planned for radiotherapy for metastatic spine disease (September 2020–July 2024). Leveraging vertebra-level expert annotations, we introduced an unconventional U-Net-based strategy converting coarse voxel-wise predictions into vertebra-level lesion classifications. The final dataset comprised 2,125 vertebrae across four classes (no lesion, osteolytic, osteoblastic, mixed), split into a 3-fold cross-validation set and an independent holdout test set. Model performance was benchmarked against a DenseNet121 baseline and a musculoskeletal radiologist, with Cohen’s kappa assessing inter-rater agreement.

**Results:** The 3D model achieved an ensemble accuracy of 84.7%, outperforming DenseNet121 (72.1%), with substantial gains in F1 score, precision, and balanced accuracy. It showed high concordance with the radiologist (Cohen’s kappa = 0.76) and comparable sensitivity and specificity across all lesion subtypes. We found both models and the radiologist to struggle with osteolytic lesions, reflecting the difficulty of distinguishing this class from age-related changes in vertebral bone density and architecture caused by benign bone lesions, age-related systemic skeletal disorders and cancer treatments.

**Conclusions:** 3D-CNNs trained with vertebra-level labels can accurately and reliably classify vertebral metastatic lesion quality from CT scans, offering a scalable path toward automated characterization of metastatic spine disease to support clinical decision-making and large-scale radiomics research.

## 1. INTRODUCTION

Bone lesion quality classification is central to fracture risk assessment in cancer patients with metastatic vertebral body disease, informing both management ^(1, 2)^ and treatment strategies ^(3)^. On X-ray or CT imaging, bone metastases are categorized as osteolytic, osteoblastic, or mixed ^(4)^. However, classification remains highly subjective ^(5)^, with poor to moderate agreement among musculoskeletal radiology and spine oncology specialists ^(6)^, particularly for mixed lesions, limiting reliable inferences about pathologic fracture risk, whether therapy is indicated, and if so, whether patients would benefit most from surgery, vertebral augmentation, or radiation.

Early image-based and machine learning approaches, including watershed algorithms with support vector machines ^(7)^ and Random Forest classifiers ^(8)^, enable bone metastases detection and osteolytic /osteoblastic differentiation ^(9)^. Although physically interpretable, these methods rely on manual feature engineering, are constrained by minimum detectable lesion size thresholds (0.3–0.5 cm), and degrade in performance when lesions are ill-defined or morphologically atypical. Deep learning (DL) methods have since been increasingly applied to classify spinal bone metastases from CT ^(10)^ or MRI ^(11)^. Two-dimensional DL models have achieved high accuracy for healthy versus lesioned vertebra classification from CT ^(10, 12)^, with CNN-based classifiers applied to regions of interest, enabling identification of specific osteolytic ^(13)^ or osteoblastic ^(14)^ lesions. Nevertheless, 2D architectures are inherently susceptible to information loss, as a single imaging plane may not fully capture lesion extent or morphology. The use of 3D U-Net architectures ^(15, 16)^ further improved classification accuracy for osteoblastic and osteolytic lesions, with performance surpassing that of expert assessors for both lesion types ^(16)^. Transfer learning approaches ^(17)^ have further improved classification across all four classes (osteoblastic, osteolytic, mixed, and no lesion) relative to a standard 2D DenseNet121 model. Despite these advances, no model has yet achieved robust, clinical-grade performance for comprehensive four-class bone metastases characterization ^(15)^.

Our objective was to evaluate a novel 3D CNN framework for vertebral bone metastases classification that integrates vertebra-level annotations with a voxel-based deep learning architecture, enhancing lesion classification performance beyond conventional 2D methods. We benchmarked our model against an existing CNN architecture and compared its performance directly against an expert radiologist.

## 2. MATERIALS AND METHODS

### 2.1 Dataset: Cancer study participants

The study cohort comprised 151 patients with vertebral body metastases who received radiation therapy at Dana Farber Cancer Institute or Brigham & Women’s Hospital (BWH) between September 2020 and July 2024 ^(18)^. Table 1 details the patients’ demographics. All patients in this study had previously consented to the Broadband biorepository research project (MGB IRB 2016P001582). Study inclusion criteria were 1) presence of at least one histologically or cytologically documented stage IV bone metastases and radiographic (CT or bone scan) evidence of bone metastases in the thoracic, thoracolumbar or lumbar spine, and 2) Karnofsky Performance Status ^(19)^ > 70. Patients were excluded if they had: 1) bone metabolism diseases (Paget, Cushing, untreated hyperthyroidism, or hyperprolactinemia); or 2) previous radiotherapy (< 6 months), surgery, or vertebral augmentation at the site of radiation or adjacent levels.

**Table 1.** Demographic characteristics of the patients included in the study.

| Characteristics | Male (n=100) |  | Female (n=50) |  |
| --- | --- | --- | --- | --- |
|  | Mean (SD) | Range | Mean (SD) | Range |
| Age (years) | 66.9 (10.4) | 34 – 87 | 60.7 (13.5) | 26 – 89 |
| Height (cm) | 176.4 (7.0) | 157.5 – 199.8 | 160.9 (7.5) | 144.5 – 175.3 |
| Body mass (kg) | 87.2 (16.4) | 55.8 – 145.3 | 63.1 (14.4) | 42.7 – 115.3 |
| BMI (kg/m<sup>2</sup>) | 28.0 (4.7) | 19.1 – 47.4 | 24.5 (5.6) | 15.4 – 43.8 |

SD: standard deviation. \* The data presented does not include data for one patient for which we did not have demographic data.

### 2.2 CT Imaging protocol

Patients were simulated for radiation treatment by our study attending physicians at the Radiation Oncology Department, BWH (Appendix A.1), using the Siemens SOMATOM Confidence (Siemens Healthcare GmbH, Erlangen, Germany) or GE Lightspeed (General Electric Medical System, Waukesha, WI) CT scanners. Simulation scan parameters are detailed in Table 2.

**Table 2.** NIH cancer study X-ray Computer tomography imaging protocol parameters.

| Radiotherapy | CT scanner |  |  |  |
| --- | --- | --- | --- | --- |
|  | Siemens SOMATOM Confidence |  | GE Lightspeed |  |
| Protocol parameters | SBRT | All Others | SBRT | All Others |
| Tube voltage (kVp) | 120 | 120 | 120 | 120 |
| Tube current (mA) | Variable | Variable | 300 | 300 |
| Field of View (FOV) | A, B | A, B | A, B | A, B |
| Slice Thickness | 0.5mm | 1.5mm | 1.25mm | 1.25mm |
| In-Plane Pixel Size | 0.31x0.31mm | 0.31x0.31mm | 0.31x0.31mm | 0.31x0.31mm |
| Gantry rotation | 1s | 1s | 1s | 1s |
| Gating | None | None | None | None |
| Breath Hold | None | None | None | None |

Variable: Tube current 240-300(mA); FOV: A: 16cm, B: Skin-to-Skin, depending on the patient's habitus. In-plane Pixel Size (mm): A: 0.31\*0.31, B: 0.70-0.98.

### 2.3. Preparation and annotation of ground truth metastatic bone annotations

#### I. Training annotations

The patient’s CT scans were reviewed by a neuroradiologist (DBH, > 41 years of clinical experience) and a spine biomechanician (RNA, >28 years of vertebral imaging and biomechanics experience). Each level with bone metastases was reviewed (DBH, RNA) and annotated as having a: 1) Osteolytic, 2) Osteoblastic or 3) Mixed lesion (Figure 1). With cancer being a systemic disease, the lack of clear evidence of a bone metastases on CT does not render the vertebra healthy. Hence, we termed such vertebrae as having no observed lesion on CT (NOL).

**Figure 1:**
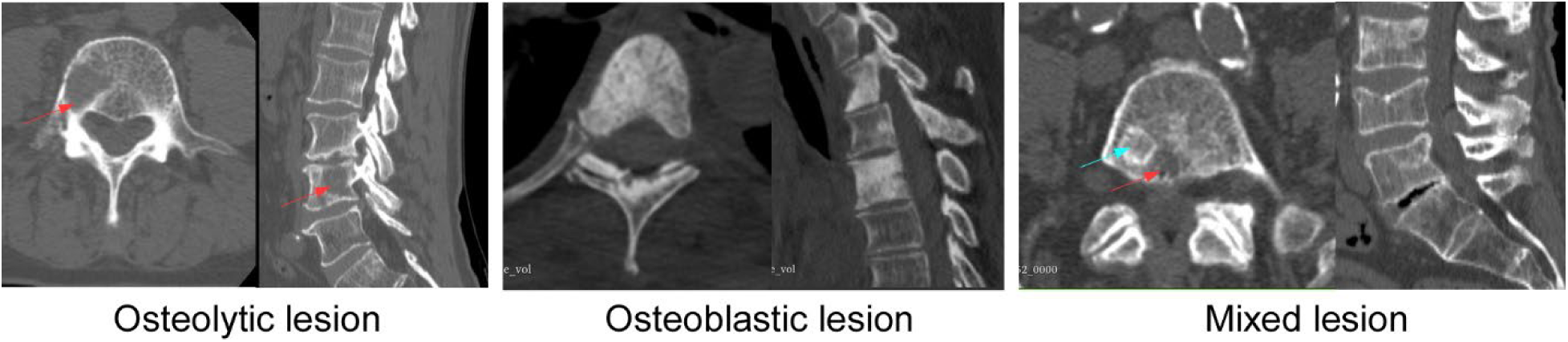
Radiographic presentation of osteolytic (marked with red arrows), osteoblastic, and mixed (osteoblastic lesion marked with a green arrow) metastatic bone lesions in cancer patients

#### II. External annotations

An external neuroradiologist (BA, >5 years of clinical experience) reviewed the patient’s CT scans from the holdout test (section 2.4.III), blind to the patient’s clinical history and demographics, with vertebral levels classified using the study terminology (section I).

### 2.4 Deep Learning-Based Algorithm

Prior to classification, an nnUNet DL model ^(20)^ was applied to segment vertebral levels, followed by precise isolation, cropping, and masking of irrelevant regions. Preprocessing included resampling to a fixed resolution, intensity clipping, histogram normalization, and standardization. Data augmentation involved random rotations and flips, with class balancing achieved through stratified sampling and per-epoch down-sampling. The dataset was split into a 5-fold cross-validation and a holdout test set (82%/18%) using per-patient stratification. Both models were trained with cross-entropy loss and the Adam optimizer for 250 epochs.

#### I. Baseline Model

Consistent with previous 2D lesion classification studies ^(10, 12)^, the baseline model employed a DenseNet121 CNN trained on cropped vertebra volumes using the MONAI framework.

#### II. New Classifier Model

In the absence of ground truth voxel-based segmentations for lesion quality, we adopted a hybrid strategy using an nnU-Net CNN to transform voxel-level predictions into vertebra-level classifications by leveraging vertebra-level lesion labels (Figure 2).

**Figure 2:**
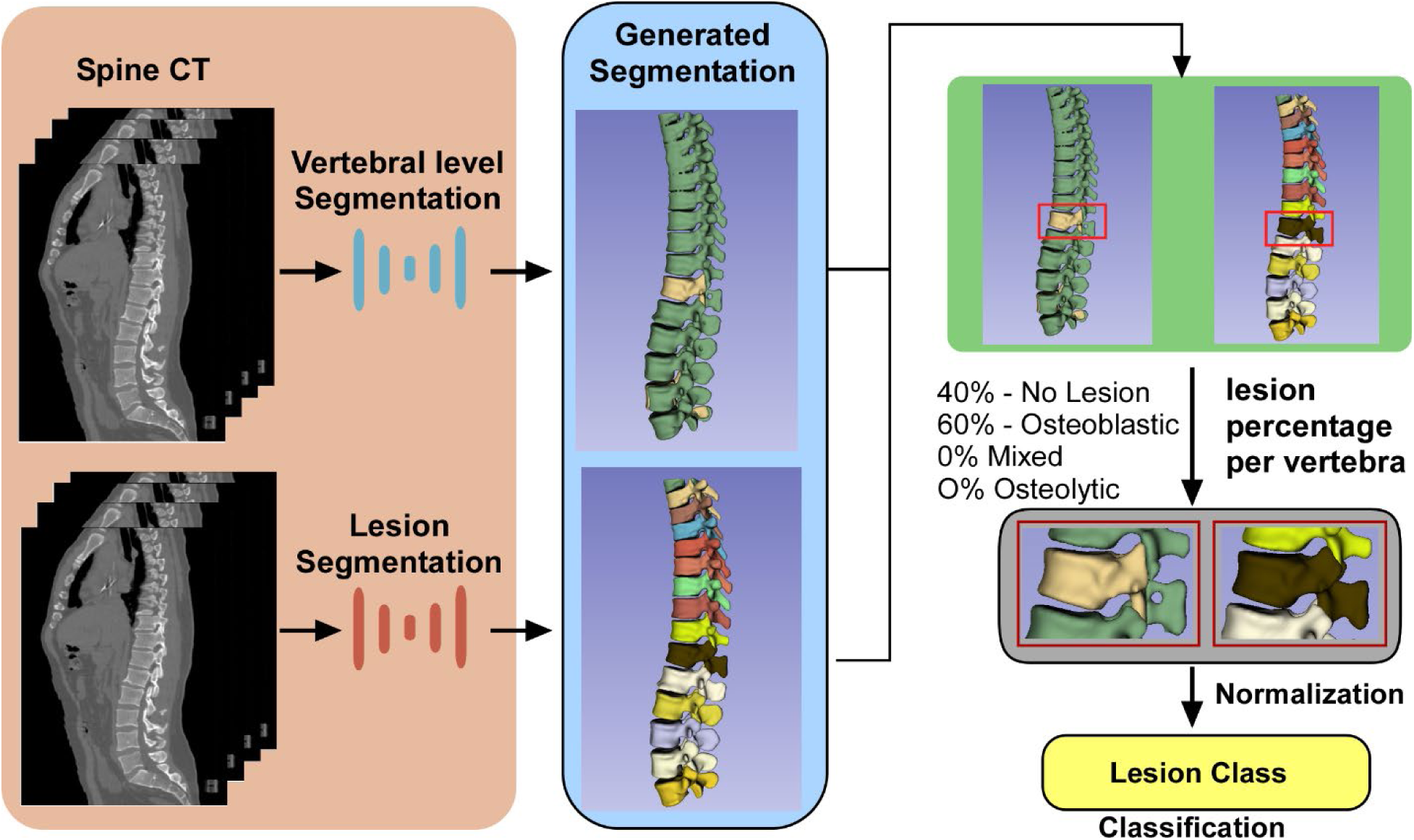
We employed a voxel-level segmentation deep neural network (nnUNet) unconventionally to transform voxel-level predictions into vertebra-level classifications.

##### Network Architecture and Design

The pipeline (Figure 2) begins with pre-processed DL ^(20)^ vertebral-level segmentations (Figure 2). Labeling every voxel within each annotated vertebra as lesional produces a coarse but complete 3D label map. Converted into a voxel-level segmentation mask, it was used to train a second segmentation model (Figure 2. Rather than interpreting its output as precise lesion boundaries, we repurposed this model as a classifier: whole-vertebra labeling is an intentional design choice that robustly captures a representative subset of lesional voxels regardless of lesion size.

To convert segmentation outputs into a per-vertebra classification score, we introduce a normalization-based aggregation scheme computing the proportion of voxels assigned to each lesion class per vertebra. Class ratios, rather than raw voxel counts, mitigate volume imbalance between lesioned and healthy tissue, particularly important for osteoblastic lesions, where non-lesioned tissue may dominate vertebral volume. Proportions are normalized using the validation set’s mean and standard deviation, with the highest normalized probability assigned as the final classification score; validation-fold distributions were aggregated into a global mean and standard deviation to ensure consistent normalization.

#### III. Training Details and Data Stratification

The dataset comprised 151 CT spines yielding 2125 vertebrae: 76% (1615) NOL, 11% (240) osteoblastic, 7% (151) osteolytic, and 6% (119) mixed. Two datasets were constructed: (1) a training set of 1738 vertebrae (82%) for 3-fold cross-validation, with one-third held out per fold, and (2) a holdout test set of 387 vertebrae (18%: 291 NOL, 59 osteoblastic, 38 osteolytic, 24 mixed). Data were stratified by lesion label and patient ID to mitigate class imbalance and prevent leakage, ensuring each spine was assigned exclusively to the training, validation, or holdout set. Model training used the 3D full-resolution nnU-Net configuration ^(21)^, with performance monitored via pseudo-Dice score, validation loss, accuracy, and EMA accuracy.

### 2.6 Statistical Analysis and Evaluation Metrics

All statistical analyses used the R computational environment ^(22)^. Weighted Cohen’s kappa ^(23)^ assessed inter-rater agreement between study annotations (ground-truth) and the external radiologist’s lesion quality classifications. For both binary detection and multiclass classification, model ensemble performance was evaluated using accuracy, balanced accuracy, recall, precision, and F1 score, each macro-averaged across classes. Area under the receiver operating characteristic curve (AUC) ^(24)^ was calculated per lesion type. Specifically:

a. **Binary classification:** The McNemar test ^(25)^ assessed agreement between each model’s binary classification (lesion vs. no lesion) and ground-truth annotations.
b. **Lesion quality classification:** ROC curves were generated per bone lesion class using a one-vs-all strategy, with predictions ranked by the proportion of lesion-labeled voxels per vertebra. The DeLong test ^(26)^ assessed differences between resulting AUCs.
c. **Diagnostic sensitivity vs. external radiologist:** Each model’s sensitivity was compared directly to radiologist sensitivity scores across lesion classes.

## 3. RESULTS

### 3.1 External Radiologist Rating Agreement with the Study Lesion Annotation (Ground Truth)

Overall, the radiologist’s lesion classification achieved a kappa of 0.70 (SE = 0.038) against our ground truth annotation. Stratified by lesion type, agreement was highest for mixed (0.86, SE = 0.06) and osteoblastic (0.81, SE = 0.41) lesions, followed by NOL (0.63, SE = 0.04), with the lowest agreement observed for osteolytic lesions (0.28, SE = 0.12).

### 3.2 The classifier model outperforms the baseline model for lesion detection

The baseline model achieved a mean validation accuracy of 67.7% ± 1.2% for binary classification (lesion vs. no-lesion). On the holdout dataset, the ensemble achieved 74.8% accuracy, 53.2% F1 score, 48.8% recall, 58.4% precision, and a kappa of 0.36 (Table 3). The classifier model achieved a mean validation accuracy of 84.5% ± 0.4%, and on the holdout dataset attained 85.2% accuracy, 69.4% F1 score, 57.1% recall, 88.5% precision, and a kappa of 0.62 (Table 3). The McNemar test confirmed significantly better agreement with ground-truth classification for the classifier model over the baseline (p < 0.001).

**Table 3:** Comparison of performance metrics on lesion detection between the Baseline DenseNet121 model and our proposed classifier. Best values per metric and category are in bold.

| Model | Accuracy | F1 Score | Recall | Precision | Balanced Accuracy | Cohen's kappa |
| --- | --- | --- | --- | --- | --- | --- |
| Baseline | 74.8 | 53.2 | 48.8 | 58.4 | 67.2 | 0.36 (0.28-0.45) |
| Classifier | **85.2** | **69.4** | **57.1** | **88.5** | **77.0** | 0.62 (0.54-0.71) |

Reported as Cohen's kappa (95% confidence intervals). Recall: sensitivity; Precision: positive predictive value.

### 3.3 Classifier Model Outperforms Baseline Model for Identification of Specific Lesion Classes

**A. Baseline:** The baseline model achieved a mean (SD) validation accuracy of 62.0% (±3.1%) across three cross-validation folds. On the holdout test set, the ensemble achieved 72.1% accuracy, an F1 score of 52.6%, 51.9% recall, and 54.6% precision for bone lesion quality classification (Table 4). Per-class accuracy was 85.6% for NOL, 49.1% for osteoblastic, 62.5% for mixed, and 10.5% for osteolytic lesions.
**B. Classifier:** Figure 3 presents radar plots summarizing the improvement in classifier performance in comparison to the baseline model for each lesion subtype and evaluation metric, with detailed comparison provided in Table 4. The classifier model achieved a mean (SD) validation accuracy of 82.4% (±0.8%), representing a 32.9% improvement over the baseline. On the holdout test set, the classifier outperformed the baseline across all metrics: accuracy (+12.6%), F1 score (+17.1%), kappa (+26.4%), recall (+15.2%), and precision (+24.7%) (Table 4).

Per-class, the classifier demonstrated higher accuracy for osteoblastic (+20.4%), mixed (+29.2%), and NOL (+11.3%) lesions, with no improvement observed for osteolytic lesions (0.0%) Figure 5. Application of the DeLong test (26) showed the improvements in prediction were significant for Mixed (p=0.0230) and osteoblastic (p<0.0001) lesion types. Table A.1 provides a detailed comparison of performance metrics between the Baseline DenseNet121 and Classifier models. Best values per metric and category are in bold.

**Figure 3:**
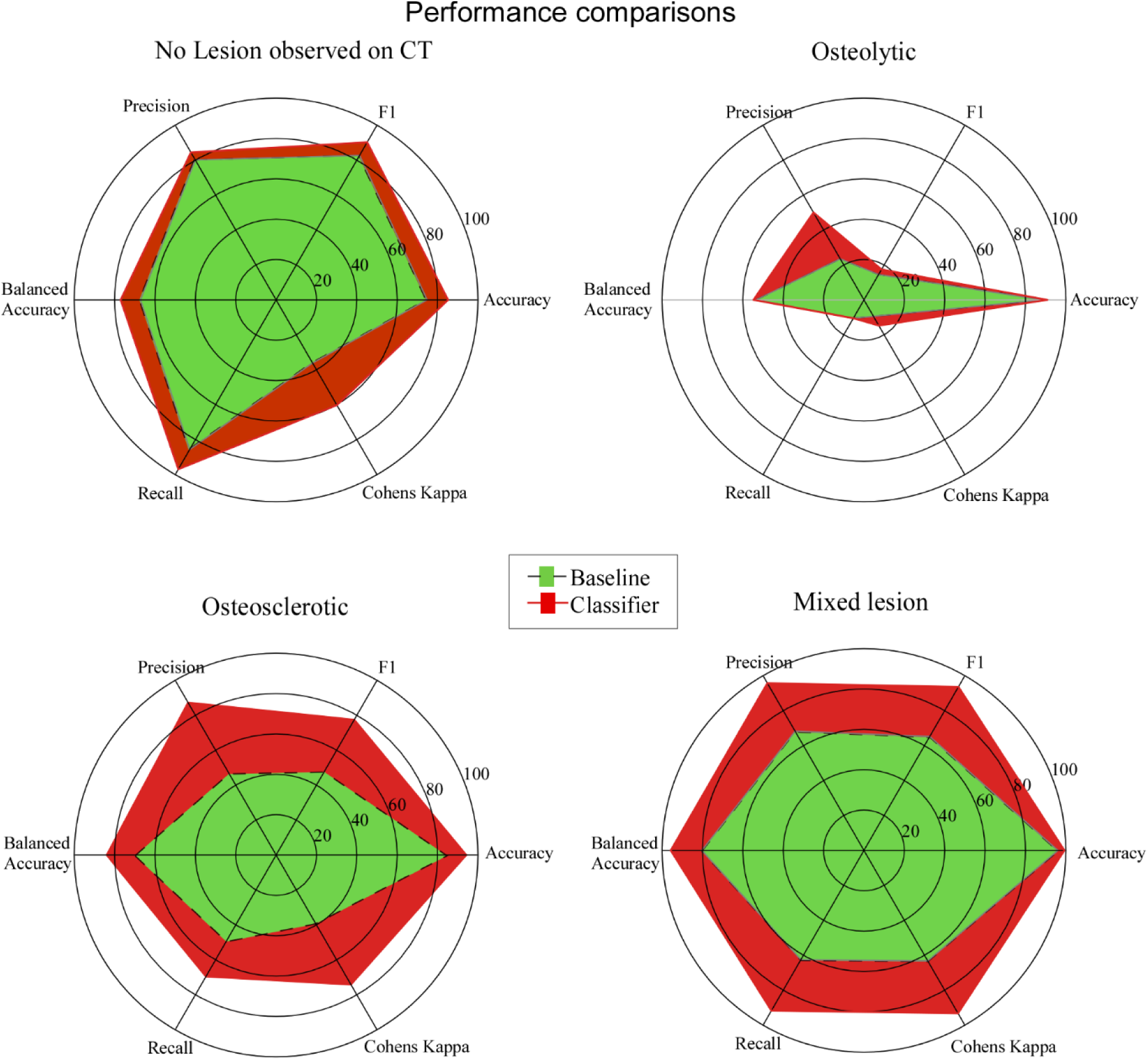
Radar plots show the classifier consistently outperforms the baseline across all metrics and lesion subtypes, with the largest gains in accuracy, F1, and Cohen’s kappa for mixed and osteoblastic lesions, indicating stronger predictive performance and better agreement with expert annotation. For osteolytic lesions, accuracy and balanced accuracy improved despite lower overall scores. These results indicate that the model generalizes well across lesion types and yields more reliable predictions than the baseline.

**Figure 4:**
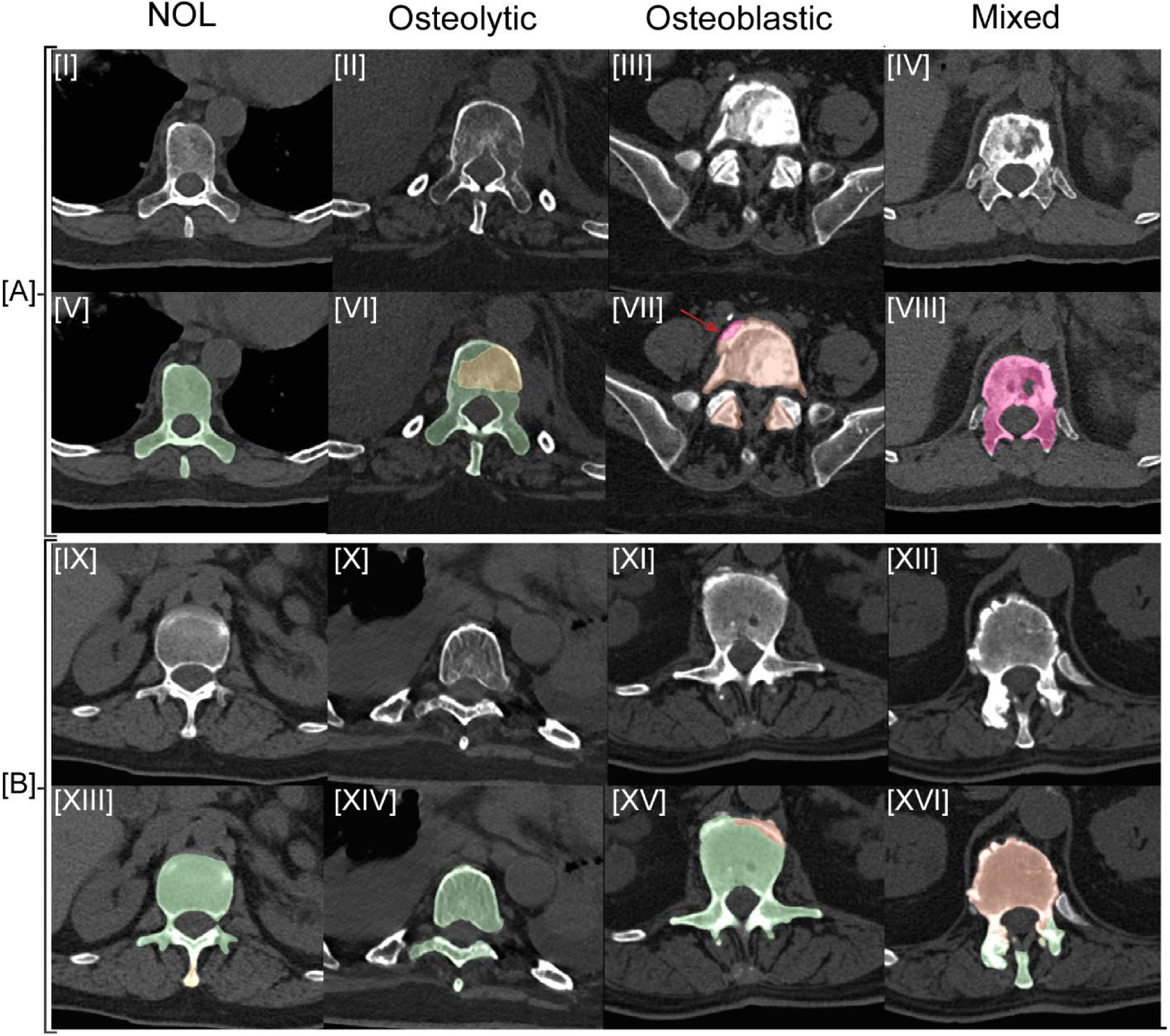
Sample correct [A] and incorrect [B] classifier model predictions for NOL, osteolytic, osteoblastic, and mixed lesions. CT images for the four lesion classes [I–IV] are shown with corresponding classifier prediction maps [V–VIII]. The red arrow in image VII highlights a vertebral osteophyte incorrectly classified as a mixed lesion. Incorrect classifier predictions for each lesion class are shown in [XIII–XVI] with corresponding source CT data [IX–XII].

**Figure 5:**
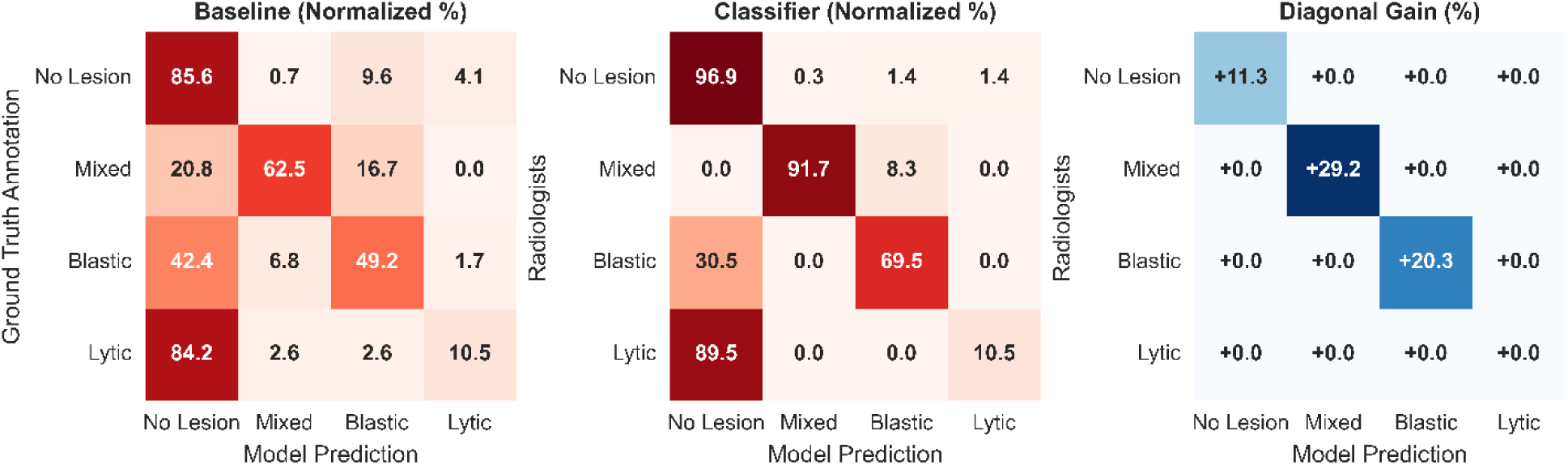
Normalized confusion matrices of both models and diagonal gain.

**Table 4:** Comparison of performance metrics between the Baseline DenseNet121 model and our proposed classifier. Best values per metric and category are in bold.

| Metric / Class | Baseline (%) | Classifier (%) | Difference (%) |
| --- | --- | --- | --- |
| Overall Metrics |  |  |  |
| Accuracy | 72.1 | **84.7** | +12.6 |
| F1 Score | 52.6 | **69.7** | +17.1 |
| Precision | 54.6 | **79.3** | +24.7 |
| Recall | 51.9 | **67.1** | +15.2 |
| Cohen's kappa | 36.3 | **62.3** | +26.4 |
| Class-Specific Accuracy |  |  |  |
| NOL | 85.6 | **96.9** | +11.3 |
| Osteoblastic | 49.1 | **69.5** | +20.4 |
| Mixed | 62.5 | **91.7** | +29.2 |
| Osteolytic | 10.5 | **10.5** | 0.0 |

### 3.4 Classifier Model Performance vs. That of a Clinical Radiologist Was Comparable to Inter-rater Agreement Among Two Radiologists

Overall classification accuracy was 84.7% for the classifier and 87.1% for the radiologist. Direct inter-rater agreement between the radiologist and the computational model yielded a kappa of 0.699 (SE = 0.038), indicating substantial concordance. Figure 6 presents ROC curves comparing baseline, classifier, and radiologist (red dot) performance by lesion subtype. The classifier outperformed the radiologist for “Mixed” lesions, matched radiologist performance for “Osteoblastic” lesions, and underperformed for “No Lesion” vertebrae. Both the model and radiologist struggled with “ Osteolytic” lesion classification (Figure 6).

**Figure 6:**
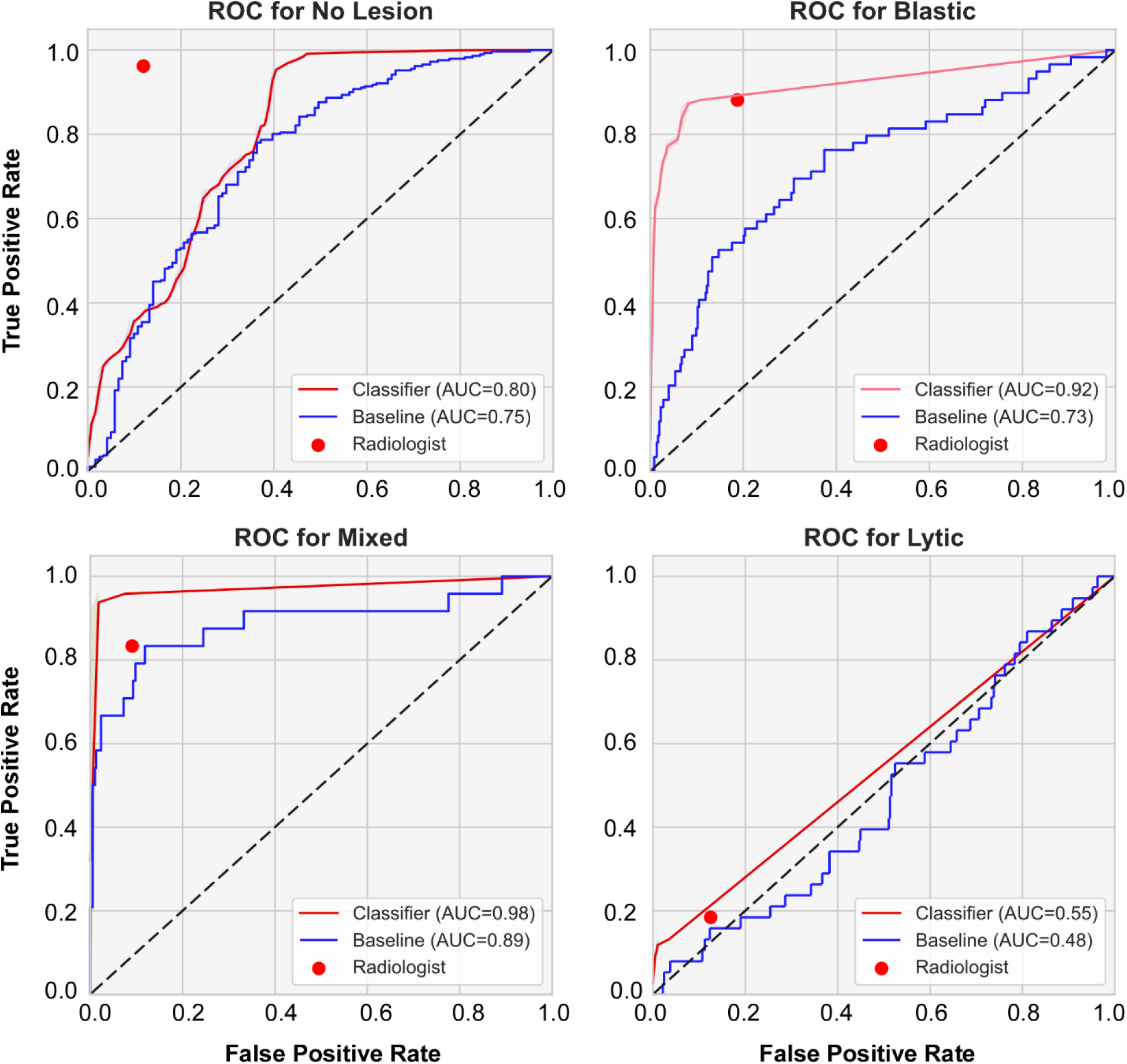
ROC curves comparing classifier, baseline model, and radiologist performance for classifying vertebral lesions by subtype.

Analysis of agreement supported these findings: the model showed higher agreement with ground-truth annotations for “Mixed” lesions (kappa = 0.93, SE = 0.04) but lower agreement for “ Osteoblastic” lesions (kappa = 0.74, SE = 0.05) compared to the radiologist (kappa = 0.81, SE = 0.41). Agreement for “No Lesion” was moderate and closely matched between the radiologist (kappa = 0.63, SE = 0.04) and the model (kappa = 0.60, SE = 0.05). “Osteolytic” lesions showed the weakest agreement for both the radiologist (kappa = 0.28, SE = 0.12) and the model (kappa = 0.15, SE = 0.13).

Lesion-level performance metrics further quantified model-radiologist agreement. Across all lesions, the ensemble achieved an F1 score of 0.77, an accuracy of 0.92, and a Cohen’s kappa of 0.76 when compared directly with the radiologist, indicating strong concordance and robust agreement with human interpretation.

## 4. DISCUSSION

Trained on coarse vertebra-level annotations, our voxel-based 3D approach demonstrated significantly higher accuracy than the 2D DenseNet121 baseline for identifying lesioned versus vertebrae with no observed lesions, and for differentiating lesion quality (osteolytic, osteoblastic, mixed). Comparison with an experienced musculoskeletal radiologist revealed substantial inter-rater agreement (Cohen’s kappa = 0.76), demonstrating that a 3D-CNN trained on vertebra-level labels can accurately characterize spinal metastasis quality from CT data, matching radiologist performance, a scalable, automated approach supporting large-scale radiomics research and clinical decision support in metastatic spine disease. Our findings showed both models and the external radiologist struggled in classifying osteolytic lesions, highlighting the difficulty of distinguishing such lesions from age-related changes in vertebral bone density and architecture caused by the features of benign bone lesions and systemic skeletal disorders, such as osteoporosis ^(27)^ or cancer-related treatments ^(28)^.

### 4.1 Detection of Spinal Metastases Versus Normal Vertebrae

The reliable classification of bone metastases, a significant clinical challenge in advanced cancer patients ^(29, 30)^, is crucial for diagnosis, management, and treatment to maintain or improve patient quality of life and life expectancy ^(1)^. Evaluated in 151 cancer patients with metastatic spine disease, the baseline DenseNet121 CNN classifier achieved 74.8% accuracy, 48.8% recall, 53.2% F1-score, and 58.4% precision. Although not directly comparable to prior single-lesion-type models, this is consistent with Chmelik et al. ^(16)^ and others ^(31, 32)^, and somewhat lower than models targeting osteoblastic (79-83%) ^(7, 9)^ or osteolytic (88%) ^(9)^ lesions alone, reflecting the greater difficulty of predicting across all three types. Using nnU-Net to transform voxel-level predictions into vertebra-level classifications, the newly developed DL classifier yielded marked gains in accuracy (85.2% vs. 74.8%), F1-score (69.4% vs. 53.2%), precision (88.5% vs. 58.4%), and recall (57.1% vs. 48.8%), with substantially improved Cohen’s kappa (0.62 vs. 0.36), a statistically significant difference, comparable to ^(13, 16, 33)^ or exceeding prior ^(31–33)^ studies. These improvements suggest the classifier model achieved enhanced predictive performance for malignant vertebrae on clinical CT.

### 4.2. Differential Performance Across Osteolytic, Osteoblastic and Mixed Lesion Types

Accurate differentiation of osteolytic, osteoblastic, or mixed metastatic bone lesions has direct clinical implications for predicting vertebral fracture risk and spinal instability ^(1)^. Prior CT-based classification has focused primarily on osteolytic lesions, given their higher fracture risk ^(1)^. Applied to 2D CT axial slices, machine learning ^(9)^, U-Net CNN ^(34)^, semantic segmentation ^(12)^, and transfer-learning ^(13)^ models reported 74-88% sensitivity for osteolytic lesion detection, while osteoblastic-specific models reported a dice similarity coefficient (DSC) of 0.74-0.83 ^(16, 35)^, a disparity similarly noted by Chang et al. ^(34, 35)^ in the same population. Extending lesion subtype prediction to 3D data remains a consistent limitation of DL models ^(9, 12, 15, 34, 35)^, attributed by Edelmers et al. ^(15)^ to osteoblastic lesions’ higher heterogeneity and subtler imaging characteristics. Mixed metastases, typical of breast, lung, and prostate cancers ^(29)^, combine osteoblastic (increased trabecular thickness and mineral content ^(4)^) and osteolytic (decreased bone volume and connectivity loss ^(4)^) features, resulting in models achieving markedly poorer accuracy ^(17, 33)^ compared to either lesion type alone. To our knowledge, no published CT model has been validated for identifying all metastatic lesion types (osteolytic, osteoblastic, mixed) against normal vertebrae in a unified architecture.

Leveraging the full spatial continuity of the CT volume, this study represents a significant advancement over prior deep learning approaches largely reliant on 2D slice-based classification ^(10)^. This distinction is particularly critical for “mixed” metastases, where osteolytic and osteoblastic components are often heterogeneously distributed across the vertebral body: a 2D model viewing a single slice might capture only one component, misclassifying the lesion as purely osteolytic or osteoblastic, whereas our volumetric approach integrates features across the entire vertebra to identify the complex pathology correctly. Our nnU-Net strategy achieved 66.8% higher Cohen’s kappa, 51.5% higher precision, 30.5% higher F1-score, 17% higher recall, and 13.9% higher accuracy versus baseline, with class-specific accuracies of 96.9% for vertebrae free of metastasis, 69.5% for osteosclerotic lesions, and 91.7% for mixed lesions, improvements of 20.4, 29.2, and 11.3 percentage points respectively compared to the DenseNet121 CNN (Table 4). We attribute this gain to two factors: nnU-Net’s superior preprocessing pipeline relative to DenseNet, and our model’s ability to leverage full-spine spatial context, capturing biological dependencies, such as increased likelihood of adjacent vertebral involvement, that isolated vertebra-level predictions inherently miss.

By comparison, we found both the DenseNet121 CNN and the classifier had low accuracy (10.5%) for classifying osteolytic lesions, with the classifier showing no improvement over the DenseNet121 CNN. Although direct comparison to previous studies is limited by differences in patient populations, imaging protocols, class distributions, lesion-selection criteria, and evaluation datasets, this finding is in contrast to other studies that reported higher class-specific performance for osteolytic lesions ^(9, 12, 15, 34, 35)^. The comparable performance of both models, despite their different preprocessing pipelines, suggests that the main limitation may not be related to data preprocessing but rather to the data itself. Our cohort, consisting predominantly of prostate, lung, breast, and renal cancer patients treated with radiotherapy for metastatic spine disease, presented mostly osteolytic lesions that were small, less well defined or diffusely distributed within the body rather than well-defined osteolytic foci. Distinguishing such lesions from the features of benign bone lesions, the loss of bone mineral density and the degradation of bone architecture caused by systemic skeletal disorders, such as osteoporosis ^(27)^ or cancer-related treatments ^(28)^, and the limited image resolution offered by clinical CT data which results in a varying degree of visual similarity between vertebrae containing small lytic lesions and older vertebrae, significantly complicating this challenge. This challenge is made clear from the weak agreement between the external radiologist and the study radiologist-generated ground truth (kappa = 0.28, SE = 0.12) as well as between the external radiologist and the model (kappa = 0.15, SE = 0.13) for classifying osteolytic lesions.

### 4.2 DL Model Performance Versus Expert Musculoskeletal Radiology Reader

In our blinded reader study, the model achieved 84.7% accuracy versus the radiologist’s 87.1%, with high concordance between the two, with the ROC analysis confirming strong discrimination between subtypes (AUC 0.91). The model’s slight underperformance in identifying pure osteolytic versus osteoblastic lesions mirrors a known radiologic challenge: early osteolytic changes often present as subtle hypo-intensities easily conflated with osteopenia or osteoporosis, whereas osteoblastic lesions present as high-contrast hyperdensities. Lesion-level performance metrics further quantified model-radiologist agreement. Across all lesions, the ensemble achieved an F1 score of 0.77, an accuracy of 0.92, and a Cohen’s kappa of 0.76 when compared directly with the radiologist, indicating strong concordance and robust agreement with human interpretation.

Our study has several limitations. First, the dataset was relatively small (151 patients), a common constraint in medical deep learning that can cause overfitting and limit generalizability; although cross-validation and data augmentation helped mitigate this, performance on rare presentations remains to be fully stress-tested. We would expect that incorporating larger cohorts with higher representation of Multiple Myeloma, Renal Cell Carcinoma, and non-small cell lung cancer patients, addressing the low number of osteolytic lesions in the study, can be expected to yield models that perform better and have more balanced performance across all 4 classes. Second, the study limited the radiologist to cropped vertebral volumes, excluding clinical history and full spinal context; while this ensured a fair “pixels-only” comparison with the AI. With the radiologists generating the ground truth labels having had access to clinical data, acting as prior information common in real-world settings, might have led both the model and reader study to underestimate the osteolytic lesions. Third, the model was restricted to imaging data and therefore did not account for clinically relevant variables such as patient age, primary cancer type, treatment history, or other medical information. In routine clinical practice, radiologists interpret imaging findings in conjunction with these contextual factors, which may help distinguish metastatic lesions from age-related or treatment-related skeletal changes. Future work could therefore investigate multimodal approaches that combine imaging features with textual or tabular clinical data. In addition, leveraging pretrained foundation models may improve feature representation and lesion recognition, particularly in settings with limited training data.

## Conclusions

We have demonstrated that a 3D-CNN trained on vertebra-level labels can accurately classify spinal metastasis subtype, matching experienced radiologists’ performance, a scalable, automated method advancing large-scale radiomics research and clinical decision support in metastatic spine disease. By standardizing this process, our model could mitigate human error and variability while serving as a triage tool flagging patients at high risk for structural instability who may require prophylactic stabilization or targeted radiotherapy. Its superior performance in identifying mixed lesions is particularly relevant, as these heterogeneous lesions are often the most difficult to characterize subjectively yet represent a significant portion of the metastatic population.

Future work must focus on multi-institutional validation using diverse CT scanners and acquisition protocols, and on integrating this classification module into a comprehensive pipeline—including detection, segmentation, and fracture risk prediction- to provide a holistic decision-support system for spine oncology. Prospective studies examining the model’s “second-reader” impact on workflow efficiency and diagnostic accuracy are also warranted.

## Data Availability

All data produced in the present study are available upon reasonable request to the authors

## Acknowledgments

The authors acknowledge the financial support of the National Institute of Arthritis and Musculoskeletal and Skin Diseases (NIAMS) under award numbers R01AR075964 and 3R01AR075964-03S1. This work used Jetstream2 at Indiana University through allocation CIS230102 from the Advanced Cyberinfrastructure Coordination Ecosystem Services and Support (ACCESS) program, which the National Science Foundation supports with grants numbers #2138259, #2138286, #2138307, #2137603, and #2138296. The authors would like to acknowledge the BROADBAND Research Project at the Brigham and Women’s Hospital Department of Radiation Oncology for providing regulatory and personnel support for this project. The BROADBAND Project was partly made possible by the generous donations of Stewart Clifford, Fredric Levin, and their families

## REFERENCES

1. Weber MH, Burch S, Buckley J, Schmidt MH, Fehlings MG, Vrionis FD, et al. Instability and impending instability of the thoracolumbar spine in patients with spinal metastases: a systematic review. Int J Oncol. 2011;38(1):5–12. PubMed PMID: 21109920.

2. Fisher CG, DiPaola CP, Ryken TC, Bilsky MH, Shaffrey CI, Berven SH, et al. A novel classification system for spinal instability in neoplastic disease: an evidence-based approach and expert consensus from the Spine Oncology Study Group. Spine (Phila Pa 1976). 2010;35(22):E1221–9. doi: 10.1097/BRS.0b013e3181e16ae2. PubMed PMID: 20562730.

3. Tomita K, Kawahara N, Kobayashi T, Yoshida A, Murakami H, Akamaru T. Surgical strategy for spinal metastases. Spine (Phila Pa 1976). 2001;26(3):298–306. doi: 10.1097/00007632-200102010-00016. PubMed PMID: 11224867.

4. Bailey S, Hackney D, Vashishth D, Alkalay RN. The effects of metastatic lesion on the structural determinants of bone: Current clinical and experimental approaches. Bone. 2020;138:115159. Epub 20191121. doi: 10.1016/j.bone.2019.115159. PubMed PMID: 31759204; PubMed Central PMCID: PMC7531290.

5. Heindel W, Gubitz R, Vieth V, Weckesser M, Schober O, Schafers M. The diagnostic imaging of bone metastases. Dtsch Arztebl Int. 2014;111(44):741–7. doi: 10.3238/arztebl.2014.0741. PubMed PMID: 25412631; PubMed Central PMCID: PMC4239579.

6. Fox S, Spiess M, Hnenny L, Fourney DR. Spinal instability neoplastic score (SINS): reliability among spine fellows and resident physicians in orthopedic surgery and neurosurgery. Global Spine Journal. 2017;7(8):744–8. doi: 10.1177/2192568217697005. PubMed Central PMCID: PMC5721994

7. Burns JE, Yao J, Wiese TS, Muñoz HE, Jones EC, Summers RM. Automated Detection of Sclerotic Metastases in the Thoracolumbar Spine at CT. Radiology. 2013;268(1):69. doi: 10.1148/radiol.13121351. PubMed PMID: 23449957; PubMed Central PMCID: PMC3689444.

8. Mehta SD, Sebro R. Random forest classifiers aid in the detection of incidental osteoblastic osseous metastases in DEXA studies. Int J Comput Assist Radiol Surg. 2019;14(5):903–9. doi: 10.1007/s11548-019-01933-1.

9. Hammon M, Dankerl P, Tsymbal A, Wels M, Kelm M, May M, et al. Automatic detection of lytic and blastic thoracolumbar spine metastases on computed tomography. European radiology. 2013;23(7):1862–70. doi: 10.1007/s00330-013-2774-5.

10. Camisa A, Montanari G, Testa A, Falzetti L, Avnet S, Baldini N, et al. Automated Detection of Spinal Lesions From CT Scans via Deep Transfer Learning. IEEE Access. 2024;12(12):2169–3536. doi: 10.1109/ACCESS.2024.3396999.

11. Liu H, Jiao M, Yuan Y, Ouyang H, Liu J, Li Y, et al. Benign and malignant diagnosis of spinal tumors based on deep learning and weighted fusion framework on MRI. Insights Imaging. 2022;13(1):1–11. doi: 10.1186/s13244-022-01227-2.

12. Motohashi M, Funauchi Y, Adachi T, Fujioka T, Otaka N, Kamiko Y. A new deep learning algorithm for detecting spinal metastases on computed tomography images. Spine 2024;49(6):390–7. doi: 10.1097/BRS.0000000000004889. PubMed PMID: 38084012; PubMed Central PMCID: PMC10898548

13. Koike Y, Yui M, Nakamura S, Yoshida A, Takegawa H, Anetai Y. Artificial intelligence-aided lytic spinal bone metastasis classification on CT scans. International Journal of Computer Assisted Radiology and Surgery. 2023;18(10):1867–74. doi: 10.1007/s11548-023-02880-8.

14. Roth HR, Yao J, Lu L, Stieger J, Burns JE, Summers RM. Detection of Sclerotic Spine Metastases via Random Aggregation of Deep Convolutional Neural Network Classifications. Recent Advances in Computational Methods and Clinical Applications for Spine Imaging. 2015:3–12. doi: 10.1007/978-3-319-14148-0_1.

15. Edelmers E, Nikulins A, Sprudza KL, Stapulone P, Puce NS, Skrebele E. AI-assisted detection and localization of spinal metastatic lesions. Diagnostics (Basel). 2024;14(21):2458. doi: 10.3390/diagnostics14212458. PubMed PMID: 39518425; PubMed Central PMCID: PMC11545154

16. Chmelik J, Jakubicek R, Walek P, Jan J, Ourednicek P, Lambert L. Deep convolutional neural network-based segmentation and classification of difficult to define metastatic spinal lesions in 3D CT data. Medical Image Analysis. 2018;49:76–88. doi: 10.1016/j.media.2018.07.008. PubMed PMID: 30114549.

17. Peng S, Lai B, Yao G, Zhang X, Zhang Y, Wang Y-F, et al. Learning-Based Bone Quality Classification Method for Spinal Metastasis. Machine Learning in Medical Imaging. 2019:426–34. doi: 10.1007/978-3-030-32692-0_49.

18. Doyle P, Caplan S, Klinger N, Shin KY, Groff M, Dillon-Martin M, et al. Spinal Instability Neoplastic Score as a Predictor of Vertebral Fracture in Patients Undergoing Radiation Therapy for Spinal Metastases: A Single-Institution Study. Adv Radiat Oncol. 2025;10(7):101803. Epub 20250509. doi: 10.1016/j.adro.2025.101803. PubMed PMID: 40548161; PubMed Central PMCID: PMC12180991.

19. Péus D, Newcomb N, Hofer S. Appraisal of the Karnofsky Performance Status and proposal of a simple algorithmic system for its evaluation. BMC Medical Informatics and Decision Making. 2013;13(72). PubMed PMID: 23870327; PubMed Central PMCID: PMC3722041.

20. Sanhinova M, Haouchine N, Pieper SD, Wells III WM, Balboni TA, Spektor A, et al., editors. Registration of longitudinal spine CTs for monitoring lesion growth. Medical Imaging 2024: Image Processing; 2024: SPIE.

21. Isensee F, Jaeger PF, Kohl SAA, Petersen J, Maier-Hein KH. nnU-Net: a self-configuring method for deep learning-based biomedical image segmentation. Nat Methods. 2020;18(2):203–11. doi: 10.1038/s41592-020-01008-z.

22. Core T. R: A language and environment for statistical computing. R Foundation for Statistical Computing. 2023.

23. McHugh ML. Interrater reliability: the kappa statistic. Biochemia medica. 2012;22(3):276–82.

24. Rainio O, Teuho J, Klen R. Evaluation metrics and statistical tests for machine learning. Sci Rep. 2024;14(1):6086. Epub 20240313. doi: 10.1038/s41598-024-56706-x. PubMed PMID: 38480847; PubMed Central PMCID: PMC10937649.

25. Agresti A. Categorical data analysis. Wiley Google Scholar. 2002;2:1174–89.

26. DeLong ER, DeLong DM, Clarke-Pearson DL. Comparing the areas under two or more correlated receiver operating characteristic curves: a nonparametric approach. Biometrics. 1988;44(3):837–45. PubMed PMID: 3203132.

27. Ruhling S, Dittmann J, Muller T, Husseini ME, Bodden J, Hernandez Petzsche MR, et al. Sex differences and age-related changes in vertebral body volume and volumetric bone mineral density at the thoracolumbar spine using opportunistic QCT. Front Endocrinol (Lausanne). 2024;15:1352048. Epub 20240215. doi: 10.3389/fendo.2024.1352048. PubMed PMID: 38440788; PubMed Central PMCID: PMC10911120.

28. Zhu N, Ni H, Guo S, Shen YQ, Chen Q. Bone complications of cancer treatment. Cancer Treat Rev. 2024;130:102828. Epub 20240907. doi: 10.1016/j.ctrv.2024.102828. PubMed PMID: 39270364.

29. Coleman R, Croucher P, Padhani A, Clezardin P, Chow E, Fallon M, et al. Bone metastases. Nat Reviews Disease Primers 2020;6(1):83. doi: 10.1038/s41572-020-00216-3 PubMed PMID: 33060614.

30. Costa L, Badia X, Chow E, Lipton A, Wardley A. Impact of skeletal complications on patients’ quality of life, mobility, and functional independence. Support Care Cancer. 2008;16(8):879–89. Epub 20080408. doi: 10.1007/s00520-008-0418-0. PubMed PMID: 18392862.

31. Ruiz J, Mahmud M, Modasshir M, Shamim Kaiser M, for the Alzheimer’s Disease Neuroimaging I. 3D DenseNet Ensemble in 4-Way Classification of Alzheimer’s Disease. Brain Informatics. 2020:85–96. doi: 10.1007/978-3-030-59277-6_8.

32. Yao J, Burns JE, Sanoria V, Summers RM. Mixed spine metastasis detection through positron emission tomography/computed tomography synthesis and multiclassifier. J Med Invest. 2017;4(2):024504. doi: 10.1117/1.JMI.4.2.024504. PubMed PMID: 28612036; PubMed Central PMCID: PMC5460314

33. Noguchi S, Nishio M, Sakamoto R, Yakami M, Fujimoto K, Emoto Y. Deep learning-based algorithm improved radiologists’ performance in bone metastases detection on CT. European Radiology. 2022;32(11):7976–87. doi: 10.1007/s00330-022-08741-3. PubMed PMID: 35394186.

34. Chang CY, Huber FA, Yeh KJ, Buckless C, Torriani M. Utilization of a convolutional neural network for automated detection of lytic spinal lesions on body CTs. Skeletal Radiology. 2023;52:1377–84. doi: 10.1007/s00256-023-04297-3. PubMed PMID: 36651936.

35. Chang CY, Buckless C, Yeh KJ, Torriani M. Automated detection and segmentation of sclerotic spinal lesions on body CTs using a deep convolutional neural network. Skeletal Radiol. 2021;51(2):391–9. doi: 10.1007/s00256-021-03873-x. PubMed PMID: 34291325.

